# Co-occurring weight- and/or shape-motivated restriction in 5,747 adults with probable avoidant/restrictive food intake disorder

**DOI:** 10.1101/2025.07.28.25332146

**Authors:** Liv Hog, Casey M. MacDermod, Jennifer P. White, Jessica H. Baker, Jerry Guintivano, Jessica S. Johnson, Shelby N. Ortiz, Emily M. Pisetsky, Nadia Micali, Cynthia M. Bulik, Laura M. Thornton, Lisa Dinkler

## Abstract

**Objective:** According to DSM-5-TR, avoidant/restrictive food intake disorder (ARFID) cannot be diagnosed alongside anorexia nervosa (AN), bulimia nervosa (BN), or any other body image disturbance. This does not accurately reflect real-world symptomatology and recent research, indicating the potential need to revise DSM-5-TR Criteria. We investigated the co-occurrence of weight- and/or shape-motivated restriction (WSR) in adults who screened positive for ARFID, providing evidence to inform such changes.

**Methods:** The sample comprised 5,747 adults who consented to participate in the ARFID-Genes and Environment (ARFID-GEN) research study, screened positive for ARFID on the NIAS and PARDI-AR-Q, and completed the EDE-Q. We placed our participants into four groups: groups one and two screened positive for AN (ARFID-AN; n=147) or BN (ARFID-BN; n=193), group three endorsed WSR without meeting AN or BN criteria (ARFID-WSR; n=2,097), and group four endorsed ARFID symptoms only (ARFID-nWSR; n=3,310). We used generalized linear models to test group differences on the NIAS, PARDI-AR-Q and EDE-Q.

**Results:** ARFID-nWSR showed lower scores than all other groups across most ARFID dimensions on the NIAS and PARDI-AR-Q, and lower odds of meeting DSM-5-TR Criteria A1 to A4 (i.e., weight loss; nutritional deficiencies; dependence on nutritional supplements; significant interference with psychosocial functioning).

**Discussion:** These findings indicate a mixed phenotype with features of both ARFID and WSR associated with more severe ARFID symptomatology. The DSM-5-TR Criteria may not capture complex real-world symptomatology in adults with probable ARFID, potentially precluding those with the most severe symptoms from receiving accurate diagnoses and appropriate care.

## Introduction

Since avoidant/restrictive food intake disorder (ARFID) was first described in the DSM-5 in 2013 (1), research has supported its classification as a distinct disorder. At the same time, research, clinical reports, and lived experience testimonies have increasingly called into question its specific diagnostic criteria. The first of these criteria (Criterion A) requires the presence of a feeding or eating disturbance (avoidance/restriction of food quantity and/or variety) that results in at least one significant physical or psychosocial consequence (i.e., weight loss; nutritional deficiencies; dependence on nutritional supplements; significant interference with psychosocial functioning) (2). The remaining three criteria serve as exclusionary criteria ensuring that the eating disturbance (1) is not explained by food insecurity or cultural/religious practices (Criterion B); (2) is not only present during the course of anorexia nervosa (AN) or bulimia nervosa (BN) and does not co-occur with weight and/or shape disturbances (Criterion C); and (3) is not better explained by another somatic or psychiatric condition (Criterion D) (2). Additionally, ARFID is often characterized by the presence of at least one ARFID dimension (i.e., sensory-based aversion, low appetite/interest in food and eating, fear of aversive consequences of eating) (3, 4). However, these dimensions are listed only as examples of restriction motivators, not as formal diagnosis requirements. ARFID is therefore primarily a diagnosis of exclusion and mainly characterized by what it is <u>not</u> (Criteria B-D), which has proven challenging in real-life application.

In recent years, Criterion C has been especially criticized by case reports, firsthand accounts, and other evidence illustrating the co-occurrence of other eating disorder (ED) psychopathology with ARFID (5–11). Furthermore, due to societal pressures to meet certain appearance ideals, weight and/or shape concerns are common in the general population, especially in those with higher BMIs (12–15). Consequently, when identifying clinically significant weight and/or shape concern when assessing ARFID, it is important to consider whether the concern translates behaviorally into restrictive eating rather than just focusing on its mere presence. Further complicating ARFID diagnosis, individuals with ARFID may interpret items on other non-ARFID ED assessments from an ARFID perspective and therefore differently than individuals with other EDs (16). For instance, the item from the Eating Disorder Examination Questionnaire (EDE-Q) (17) “Have you tried to follow definite rules regarding your eating (for example, a calorie limit) in order to influence your shape or weight (whether or not you have succeeded)?” may be scored high by individuals with ARFID and low weight who are trying to reach a healthy weight. Together, these factors create confusion in applying DSM-5-TR Criteria in both research and clinical settings which, hinders treatment development and may negatively impact patient outcomes. Clarifying the overlap and boundaries between ARFID and other EDs is therefore of significant clinical importance.

The Pica, ARFID, & Rumination Disorder Interview–ARFID Questionnaire (PARDI-AR-Q) (18) and the Nine-Item ARFID Screen (NIAS) (19) are commonly used self-report questionnaires to assess ARFID symptoms and produce a diagnostic prediction of ARFID. The PARDI-AR-Q identifies possible ARFID by evaluating DSM-5-TR Criterion A (i.e., weight loss, nutritional deficiencies, dependence on nutritional supplements, significant interference with psychosocial functioning) and the NIAS by evaluating the three ARFID dimensions (i.e., sensory-based aversion, low appetite/interest in food and eating, fear of aversive consequences of eating) using dimension cutoffs. Positive diagnostic predictions from either of these questionnaires have been used in previous studies to identify individuals with ARFID (20–22). The EDE-Q (17) evaluates non-ARFID ED behaviors and cognitions. It has been used in combination with the PARDI-AR-Q and NIAS to exclude individuals from ARFID case designations based on Criterion C (20, 23, 24). To identify those to be excluded, researchers have used the EDE-Q global score (e.g., < 2.3 or < 4) or ED behaviors (e.g., binge eating, self-induced vomiting, laxative use; items 13-17) (20, 23). These applications mirror interpretations of Criterion C as a broad exclusion of almost any other co-occurring ED pathology.

However, considerable uncertainty exists around how strict exclusion criteria based on the presence of other ED features should be, because recent research utilizing a combination of these questionnaires reveals significant overlap between symptoms of ARFID and other EDs. Individuals with AN, BN, and binge-eating disorder (BED) present with higher NIAS subscale scores than those without any ED (23, 25, 26). The reverse is also true: research and lived-experience reports reveal that some individuals with ARFID endorse weight- and/or shape-related ED cognitions and behaviors such as body dissatisfaction, a desire for thinness, and binge eating (5–7, 9, 10). Additionally, in a sample of outpatients with EDs, Abber et al. (5) identified four latent profiles: two characterized by ARFID symptoms only, one by ED symptoms only, and one by both. These findings highlight the validity of an ARFID-only phenotype but also provide evidence for a mixed phenotype characterized by ARFID and other ED symptoms. Research on how individuals with ARFID respond to the EDE-Q is scarce (27). Nevertheless, global score cutoffs have been used in research to exclude individuals being assessed for ARFID (20, 23). This overlap of ARFID- and other ED symptoms highlights the lack of clarity in how the DSM-5-TR distinguishes ARFID from other EDs, significantly contributing to confusion in applying its diagnostic criteria.

Based on this evidence, Zickgraf et al. (28), using both research and clinical evidence, proposed changes to DSM-5-TR Criteria that address the misalignment of published diagnostic criteria. Their goal is to simplify differential diagnosis in the context of both non-ARFID EDs and other conditions. The authors advocate for a positive and functional definition of ARFID (i.e., stating what ARFID is instead of what it is not) by calling for the formal incorporation of the three ARFID dimensions into the diagnostic criteria as the defining characteristic of ARFID (proposed Criterion A). Because the physical and psychosocial consequences listed in DSM-5-TR Criterion A fully overlap with those seen in other EDs, the authors suggest their inclusion as Criterion B (proposed Criterion B). They also argue for allowing an ARFID diagnosis in the presence of other ED psychopathology if <u>both</u> ARFID dimensions <u>and</u> significant weight and/or shape concerns motivate disordered eating behaviors that result in the consequences listed in proposed Criterion B.

To advance this discussion and provide empirical data on the proposed changes to the DSM-5-TR Criteria, we examined the co-occurrence of ARFID and other ED symptoms using NIAS, PARDI-AR-Q, and EDE-Q data from individuals who consented to participate in the ARFID- Genes and Environment (ARFID-GEN) research study (20), and screened positive for ARFID DSM-5-TR Criterion A on the PARDI-AR-Q and scored above cutoff on at least one of the ARFID dimensions on NIAS (i.e., had probable ARFID). Specifically, we

**a)** Used the EDE-Q to distinguish between those who may be excluded from a DSM-5-TR ARFID diagnosis based on Criterion C and those who would likely meet Criterion C. As a result, we identified four groups: (1) probable ARFID with probable AN (ARFID-AN), (2) probable ARFID with probable BN (ARFID-BN), (3) probable ARFID with weight- and/or shape-motivated restriction (ARFID-WSR), and (4) probable ARFID without weight- and/or shape-motivated restriction (ARFID-nWSR; i.e., those who meet DSM-5-TR Criterion C).
**b)** Described demographic variables, BMI, ARFID symptoms, and other ED symptoms for each of the four groups and evaluated differences in these measures between ARFID-nWSR and the other groups.

## Methods

### Participants

The current data are from ARFID-GEN, a large-scale genetic research study that examines the phenotypic and genotypic architecture of ARFID (20). It was funded by the National Institute of Mental Health, registered on ClinicalTrials.gov (NCT05605067), and approved by the UNC-Chapel Hill Institutional Review Board. Between January 2023 and May 2024, participants were recruited, provided informed consent and participated remotely by providing a saliva sample for DNA extraction and answering online questionnaires about current eating behaviors, health history, and psychiatric comorbidities. Participants were recruited through social media, healthcare providers, and advocacy organizations. Recruitment materials described ARFID symptoms to target individuals who recognized these experiences in themselves. Enrollment criteria for ARFID-GEN included fluency in English and a mailing address in the United States. Eligibility was determined using the NIAS, PARDI-AR-Q, and EDE-Q. Independent of their inclusion in ARFID-GEN, the current study comprised all individuals who met ARFID criteria on <u>both</u> the NIAS <u>and</u> the PARDI-AR-Q (probable ARFID), and who completed the EDE-Q. We chose to include only those who met criteria on both ARFID measures to ensure that all participants met DSM-5-TR Criterion A, but also met Criteria A and B as proposed by Zickgraf et al. (28).

### Measures

#### Nine-Item ARFID Screen (NIAS)

The NIAS is a 9-item self-report questionnaire assessing the current endorsement of ARFID dimensions: picky eating (*NIAS Picky eating*); low appetite (*NIAS Low appetite*); and fear of aversive consequences of eating (*NIAS Fear*) (19). Each item is scored on a 6-point Likert scale (0–5) from “Strongly disagree” to “Strongly agree.” Scores for each ARFID dimension range from 0 to 15 and are calculated by summing the respective items. The ARFID dimension scores can be summed to generate a total score (*NIAS Total Score*), which ranges from 0 to 45. To meet screening criteria for ARFID, individuals must score > 8 on *NIAS Low appetite* or > 9 on *NIAS Picky eating* or *NIAS Fear* (23). In the current study, all subscales demonstrated acceptable to excellent internal consistency, with Cronbach’s alphas of α = .77 (*NIAS Total Score*), α = .83 (*NIAS Picky Eating*), α = .87 (*NIAS Low appetite*), and α = .90 (*NIAS Fear*).

#### PARDI-ARFID-Questionnaire (PARDI-AR-Q)

The PARDI-AR-Q is a 32-item self-report questionnaire assessing ARFID symptomatology over the past one to three months and produces an ARFID diagnostic prediction based on DSM-5-TR Criterion A (i.e., A1: weight loss; A2: nutritional deficiencies; A3: dependence on nutritional supplements; A4: significant interference with psychosocial functioning), three subscale scores (*PARDI-AR-Q Sensory*, *PARDI-AR-Q Lack of interest*, *PARDI-AR-Q Concern*) corresponding to the ARFID dimensions, and a severity of impact score (*PARDI-AR-Q Severity*) (18). Subscale scores are calculated as the mean of three items rated on a 7-point Likert scale (0-6). Severity is calculated as the mean of two items rated on a 7-point Likert scale (0-6) that assess social impairment due to eating problems. Published criteria for an ARFID diagnostic prediction require an eating problem that results in at least one of the physical or psychosocial consequences outlined in DSM-5-TR Criterion A (18). In the current study, all subscales demonstrated acceptable to good internal consistency, with Cronbach’s alphas of α = .81 (*PARDI-AR-Q Sensory*), α = .76 (*PARDI-AR-Q Lack of interest*), α = .88 (*PARDI-AR-Q Concern*), and α = .81 (*PARDI-AR-Q Severity*).

#### Eating Disorder Examination Questionnaire (EDE-Q)

The EDE-Q version 6.0 (17) is a 28-item self-report questionnaire evaluating ED cognitions and behaviors in the last 28 days. Subscale scores (*Restraint*, *Eating concern*, *Weight concern*, and *Shape concern*) are calculated as the mean of items rated on a 7-point Likert scale (0-6). The mean of these subscales produces a *Global score* (0-6). Additional items assess the frequency of binge-eating episodes and compensatory behaviors (i.e., self-induced vomiting, laxative use, and driven exercise; items 13-18). The items assessing binge-eating episodes, self-induced vomiting, and laxative use were dichotomized to reflect the presence/absence of the respective behaviors. In the current study, all subscales demonstrated good to excellent internal consistency, with Cronbach’s alphas of α = .92 (*Global Score*), α = .93 (*Restraint*), α = .83 (*Eating concern*), α = .87 (*Weight concern*), and α = .92 (*Shape concern*).

### Group definitions

Figure 1 presents a flowchart depicting the categorization of our study groups. To differentiate participants who would likely meet DSM-5-TR Criterion C from those who would not, we first applied EDE-Q diagnostic algorithms for AN and BN suggested by Berg et al. (29). Participants who met criteria for AN or BN were classified as probable ARFID with probable AN (*ARFID-AN;* n=147) or probable ARFID with probable BN (*ARFID-BN;* n=193), respectively. We did not identify individuals with BED or other specified feeding or eating disorder (OSFED), as BED can co-occur with ARFID according to DSM-5-TR (2), and no diagnostic algorithm for OSFED was provided by Berg et al. (29).

**Figure 1.**
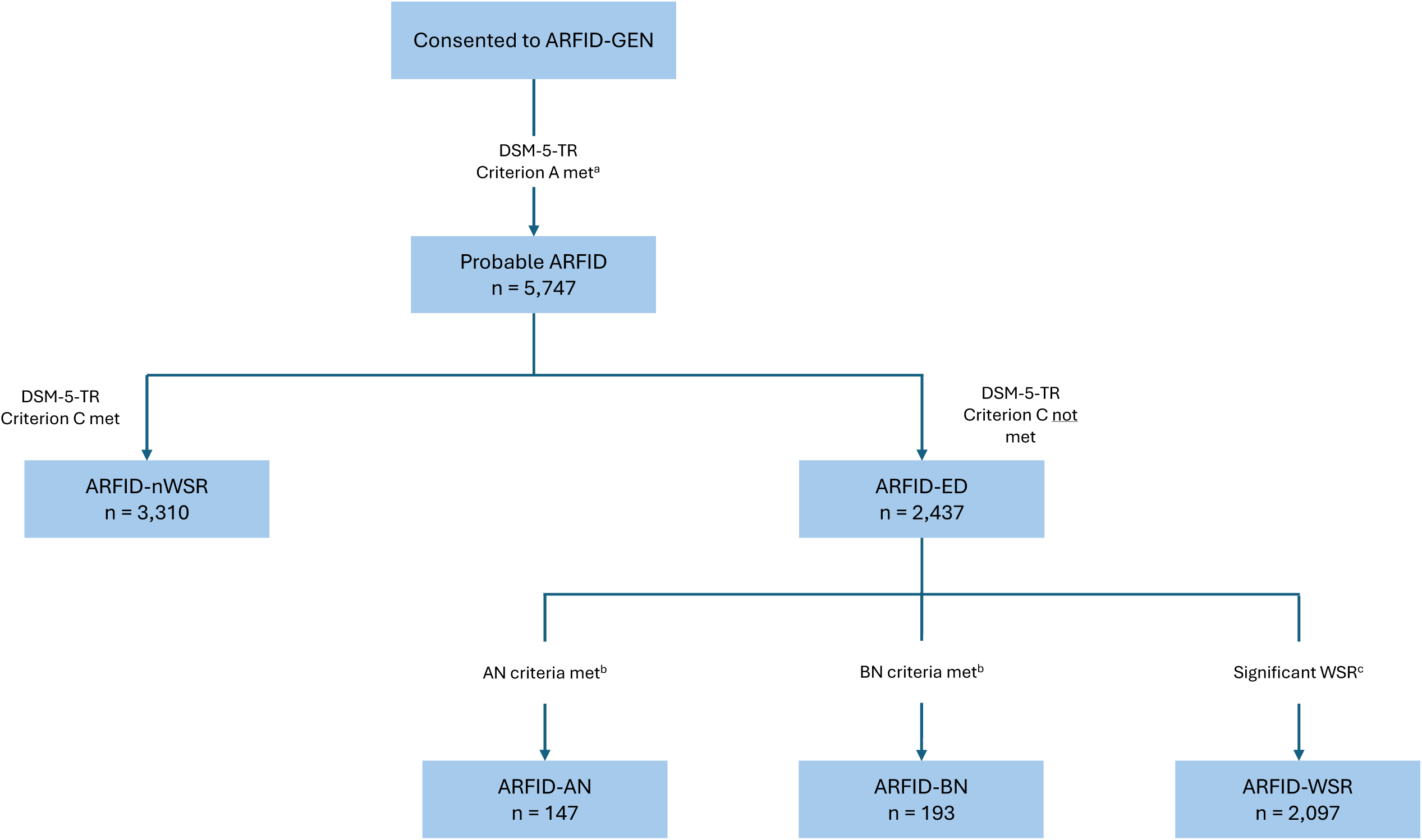
Flowchart illustrating participant group definitions. **Notes.** ARFID-GEN = ARFID-Genes and Environment research study, WSR = weight- and/or shape-motivated restriction ^a^ Positive screening on the Nine Item ARFID Screen (NIAS) and the PARDI-ARFID-Questionnaire (PARDI-AR-Q) ^b^ Using diagnostic algorithms by Berg et al. (2012) ^c^ As measured by Eating Disorder Examination Questionnaire (EDE-Q) items and cutoffs chosen by the authors to define weight- and/or shape-motivated restriction (WSR) (Table 1).

**Table 1.**
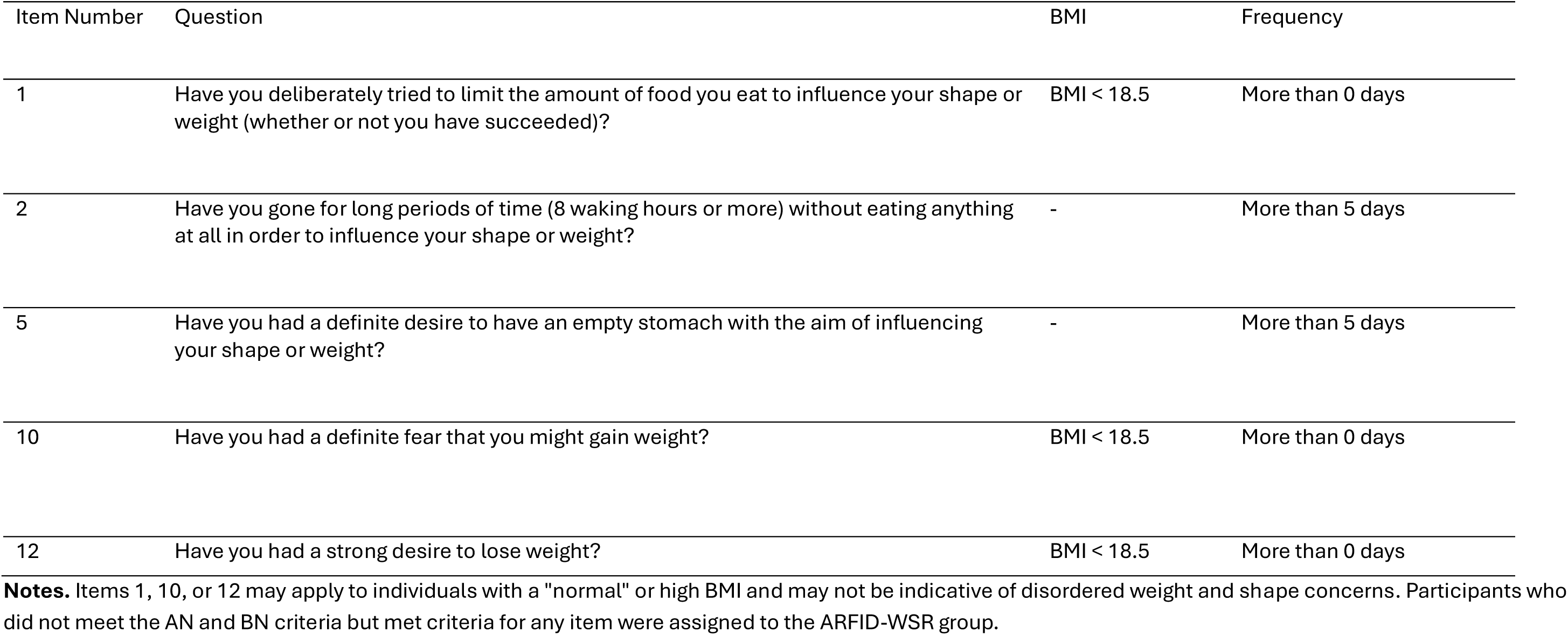
Eating Disorder Examination Questionnaire (EDE-Q; Fairburn & Beglin, 2008) items and cutoffs used to define the group of individuals with ARFID symptoms and weight- and/or shape-motivated restriction (WSR).

| Item Number | Question | BMI | Frequency |
| --- | --- | --- | --- |
| 1 | Have you deliberately tried to limit the amount of food you eat to influence your shape or weight (whether or not you have succeeded)? | BMI < 18.5 | More than 0 days |
| 2 | Have you gone for long periods of time (8 waking hours or more) without eating anything at all in order to influence your shape or weight? | - | More than 5 days |
| 5 | Have you had a definite desire to have an empty stomach with the aim of influencing your shape or weight? | - | More than 5 days |
| 10 | Have you had a definite fear that you might gain weight? | BMI < 18.5 | More than 0 days |
| 12 | Have you had a strong desire to lose weight? | BMI < 18.5 | More than 0 days |

**Notes.** Items 1, 10, or 12 may apply to individuals with a "normal" or high BMI and may not be indicative of disordered weight and shape concerns. Participants who did not meet the AN and BN criteria but met criteria for any item were assigned to the ARFID-WSR group.

Next, we identified those who, despite not having AN or BN, still may be excluded based on Criterion C. We determined whether the reported restriction could be traced back to weight or shape concerns by reviewing each question of the EDE-Q under the consideration of potential ARFID-specific interpretations and BMI (Table S1). For example, we determined that individuals at a high BM may score high on Item 1, which asks about limiting food to influence weight or shape, due to that individual following medical recommendation of weight loss as opposed to another ED cognition. Other EDE-Q items may be rated high by individuals with ARFID due to them worrying a lot about food and eating (e.g., items 7, 21) or their low weight (e.g., items 23, 24). In total, we selected five items that we judged as most suitable for identifying weight- and/or shape-motivated restriction (WSR) in people with ARFID and developed cutoff scores for each item (Table 1). Items 1, 10, and 12 were assessed only for those at low weight (BMI ≤ 18.55). Participants who exceeded cutoffs for any item were classified into the group probable ARFID with WSR (ARFID-WSR; n=2,097), whereas those who scored under cutoffs for all items were assigned to the group probable ARFID without WSR (ARFID-nWSR; n=3,310) therefore representing the only group that would likely meet DSM-5-TR Criterion C.

### Statistical analysis

Statistical analyses were conducted using SAS software, Version 9.4 (30). Descriptive statistics were calculated for the complete sample and by group for demographic variables (age, sex, race, ethnicity); BMI; BMI group (≤18.5, >18.5 and <25, ≥25 and <30, ≥30); EDE-Q subscale scores; EDE-Q global score; presence/absence of binge eating, self-induced vomiting, and laxative use; PARDI-AR-Q Criteria A1-A4; PARDI-AR-Q subscale scores; and NIAS subscale scores. Generalized linear models were applied using *proc genmod* using appropriate distributions and link functions for each outcome to test group differences; sex and age were entered as covariates into all models. Bonferroni corrections were used to adjust for multiple testing, and the threshold for significance was determined to be p < .0025 (20 tests). When omnibus tests were significant, pairwise comparisons were conducted comparing *ARFID-nWSR* to each of the other groups using Wald Chi-square tests. To ensure that potential group differences on the EDE-Q were not solely driven by items used for the group definition, we conducted a sensitivity analysis excluding those items from the EDE-Q scores. Pearson correlations were calculated among NIAS, PARDI-AR-Q, and EDE-Q subscales.

## Results

Our final analytic sample included 5,747 participants (M_age_ = 32.21 years, SD = 13.38). Most of our sample was female (89.6%), White (83.71%), and non-Hispanic (86.97%). Complete demographic information of the full sample and by group can be found in Table 2. Pearson correlations among NIAS, PARDI-AR-Q, and EDE-Q subscales are presented in Table S2.

**Table 2.** Demographic characteristics of the complete sample and by group including results of generalized linear models to test group differences.

|  |  |  | DSM-5 criterion C exclusion group (stepwise) |  |  |  | Test statistics |  |  |
| --- | --- | --- | --- | --- | --- | --- | --- | --- | --- |
| | | | Total sample<br>N = 5747 | ARFID-AN | ARFID-BN | ARFID-WSR<sup>a</sup> | ARFID-nWSR<sup>b</sup> | $\chi^2$ (df) | p value |
|  |  |  |  | N = 147 | N = 193 | N = 2097 | N = 3310 |  |  |
| **Age** |  | *M (SD)* | 32.21 (13.38) | 32.93 (13.91) | 36.67 (13.64) | 33.97 (14.25) | 30.80 (12.56) | 95.88 (3) | <.0001 |
| **Sex** | Female<sup>c</sup> | *N (%)* | 5149 (89.59) | 121 (82.31) | 154 (79.79) | 1850 (88.22) | 3024 (91.36) | 45.40 (3) | <.0001 |
| **BMI** |  | *M (SD)* | 27.64 (8.98) | 16.44 (2.27) | 29.97 (8.93) | 29.20 (9.11) | 27.01 (8.66) | 330.07 (3) | <.0001 |
| **BMI group** | ≤ 18.5 | *N (%)* | 609 (10.60) | 147 (100) | 0 | 137 (6.53) | 325 (9.82) | 1389.23 (9) | <.0001 |
|  | > 18.5 and < 25 | *N (%)* | 2129 (37.05) | 0 | 68 (35.23) | 667 (31.81) | 1394 (42.11) |  |  |
|  | ≥ 25 and < 30 | *N (%)* | 1133 (19.71) | 0 | 43 (22.28) | 468 (22.32) | 622 (18.79) |  |  |
|  | ≥ 30 | *N (%)* | 1876 (32.64) | 0 | 82 (42.49) | 825 (39.34) | 969 (29.27) |  |  |
| **Race** | White | *N (%)* | 4811 (83.71) | 126 (85.71) | 134 (69.43) | 1678 (80.02) | 2873 (86.80) | 117.20 (15) | <.0001 |
|  | Black or African American | *N (%)* | 273 (4.75) | 11 (7.48) | 29 (15.03) | 128 (6.10) | 105 (3.17) |  |  |
|  | Asian | *N (%)* | 151 (2.63) | 4 (2.72) | 9 (4.66) | 81 (3.86) | 57 (1.72) |  |  |
|  | Pacific Islander | *N (%)* | S | S | S | S | S |  |  |
|  | Native American | *N (%)* | S | S | S | S | S |  |  |
|  | More than one race | *N (%)* | 343 (5.97) | 5 (3.40) | 13 (6.74) | 138 (6.58) | 187 (5.65) |  |  |
|  | Did not answer | *N (%)* | S | S | S | S | S |  |  |
| **Ethnicity** | Hispanic | *N (%)* | 528 (9.19) | 17 (11.56) | 43 (22.28) | 192 (9.16) | 276 (8.34) | 46.94 (3) | <.0001 |
|  | Non-Hispanic | *N (%)* | 4998 (86.97) | 122 (82.99) | 137 (70.98) | 1826 (87.08) | 2913 (88.01) |  |  |
|  | Did not answer | *N (%)* | 221 (3.85) | 8 (5.44) | 13 (6.74) | 79 (3.77) | 121 (3.66) |  |  |

**Notes.** S = cell sizes were suppressed to protect privacy of participants. <sup>a</sup>Weight- and/or shape-motivated restriction defined by cutoff scores on EDE-Q items 1, 2, 5, 10, and 12 (see table 1). <sup>b</sup>Group meets DSM-5-TR Criterion A (based on NIAS and PARDI-AR-Q criteria) and Criterion C (eating problem does not occur only during the course of anorexia nervosa or bulimia nervosa and restriction not motivated by weight or shape concerns). <sup>c</sup>The remaining category comprises participants identifying as “Male” or “Other”. These groups are not presented separately due to small cell sizes, in order to protect participant privacy.

Table 3 outlines ED symptoms as measured by EDE-Q, NIAS, and PARDI-AR-Q of the full sample and of each group. Generalized linear models showed significant group differences on age, BMI, and all outcomes on EDE-Q, NIAS, and PARDI-AR-Q, except *NIAS Picky eating* (Table 3). Figure 2 presents forest plots showing mean differences (MD) for continuous outcomes and odds ratios (OR) for dichotomous outcomes. Test statistics for pairwise comparisons can be found in Tables 4 and 5 and are discussed in detail below.

**Figure 2.**
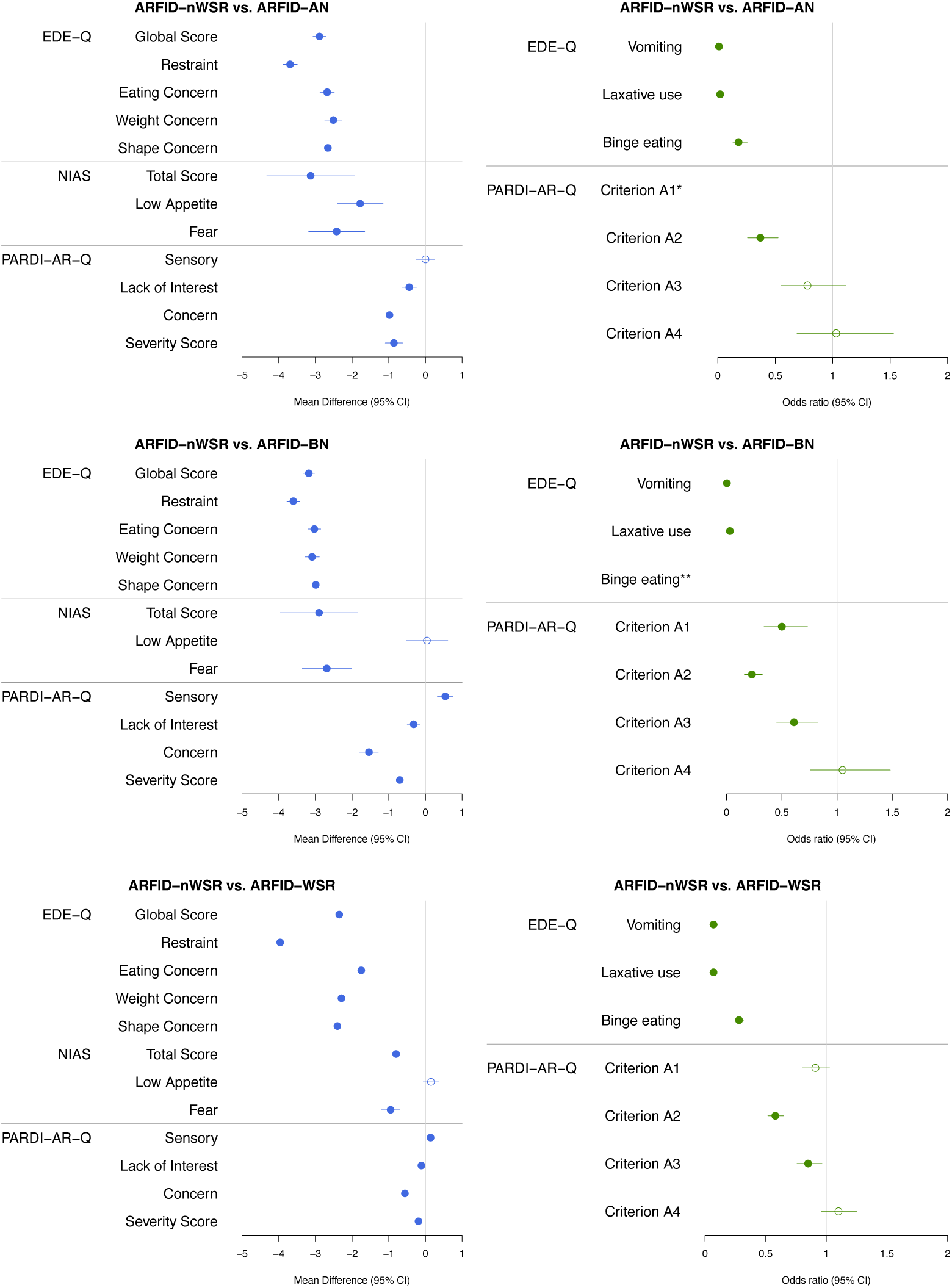
Forest plots of mean differences and odds ratios for EDE-Q, NIAS- and PARDI-AR-Q outcomes comparing ARFID-nWSR with ARFID-AN, ARFID-BN, and ARFID-WSR. **Notes.** EDE-Q = Eating Disorder Examination Questionnaire, NIAS = Nine Item ARFID Screen, PARDI-AR-Q = PARDI-ARFID-Questionnaire, PARDI-AR-Q Criteria A1-A4 = weight loss; nutritional deficiencies; dependence on nutritional supplements; significant interference with psychosocial functioning. *Pairwise comparisons not evaluated between ARFID-nWSR and ARFID-AN, since all individuals in ARFID-AN endorse Criterion A1. **Pairwise comparisons not evaluated between ARFID-nWSR and ARFID-BN, since all individuals in ARFID-BN endorse binge eating by definition.

**Table 3.**
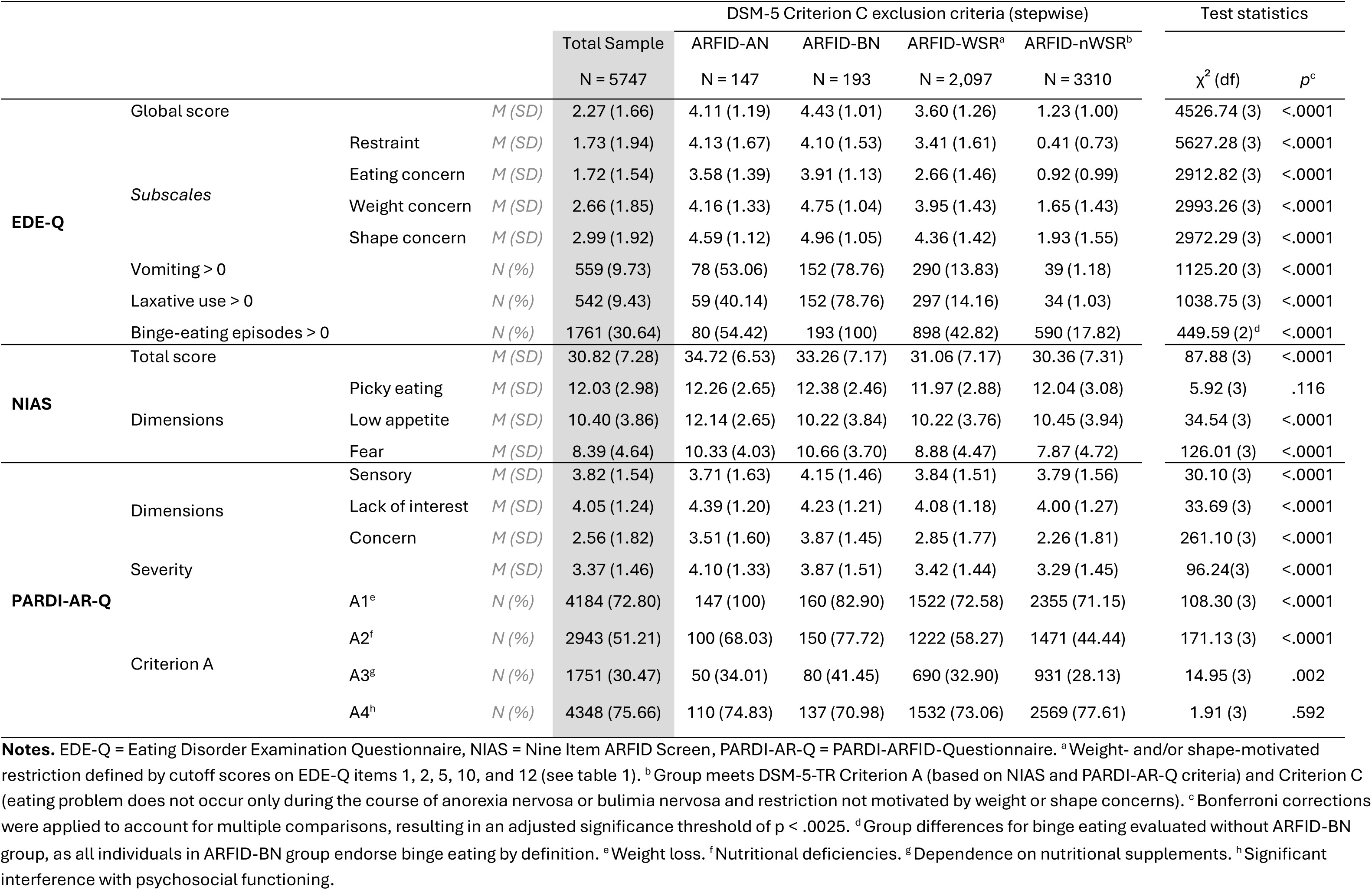
Eating disorder symptoms measured by EDE-Q, NIAS, and PARDI-AR-Q of the complete sample and by group including results of generalized linear models to tests group differences, with sex and age as covariates.

**Table 4.** Pairwise comparisons between the ARFID-nWSR and the ARFID-AN, ARFID-BN, and ARFID-WSR (referent groups) for BMI, age, and EDE-Q, NIAS, and PARDI-AR-Q scores.

|  |  | ARFID-nWSR vs. ARFID-AN |  |  | ARFID-nWSR vs. ARFID-BN |  |  | ARFID-nWSR vs. ARFID-WSR |  |  |
| --- | --- | --- | --- | --- | --- | --- | --- | --- | --- | --- |
|  |  | MD (SE) | *p* | Cohen's *d* | MD (SE) | *p* | Cohen's *d* | MD (SE) | *p* | Cohen's *d* |
| BMI |  | 10.57 (0.74) | <.0001 | 1.25 | -2.96 (0.65) | <.0001 | 0.34 | -2.19 (0.24) | <.0001 | 0.25 |
| Age |  | -2.13 (1.12) | .35 | 0.17 | -5.87 (0.98) | <.0001 | 0.47 | -3.17 (0.37) | <.0001 | 0.24 |
| **EDE-Q** | Global score | -2.89 (0.09) | <.0001 | 2.86 | -3.18 (0.08) | <.0001 | 3.20 | -2.35 (0.03) | <.0001 | 2.13 |
|  | Restraint | -3.69 (0.10) | <.0001 | 4.69 | -3.60 (0.09) | <.0001 | 4.63 | -3.96 (0.03) | <.0001 | 2.61 |
|  | Eating concern | -2.68 (0.10) | <.0001 | 2.64 | -3.03 (0.09) | <.0001 | 3.01 | -1.75 (0.03) | <.0001 | 1.46 |
|  | Weight concern | -2.51 (0.12) | <.0001 | 1.76 | -3.09 (0.10) | <.0001 | 2.20 | -2.29 (0.04) | <.0001 | 1.61 |
|  | Shape concern | -2.66 (0.12) | <.0001 | 1.73 | -2.99 (0.11) | <.0001 | 1.98 | -2.40 (0.04) | <.0001 | 1.62 |
| **NIAS** | Total score | -3.13 (0.61) | <.0001 | 0.60 | -2.90 (0.54) | <.0001 | 0.40 | -0.80 (0.20) | .003 | 0.10 |
|  | Picky eating | NE* | NE | NE | NE | NE | NE | NE | NE | NE |
|  | Low appetite | -1.78 (0.32) | <.0001 | 0.43 | 0.04 (0.29) | 1.00 | 0.60 | 0.15 (0.11) | 1.00 | 0.06 |
|  | Fear | -2.42 (0.39) | <.0001 | 0.52 | -2.69 (0.34) | <.0001 | 0.60 | -0.95 (0.13) | <.0001 | 0.22 |
| **PARDI-AR-Q** | Sensory | 0.00 (0.13) | 1.00 | 0.05 | 0.54 (0.11) | <.0001 | 0.24 | 0.14 (0.04) | .002 | 0.03 |
|  | Lack of interest | -0.44 (0.10) | <.0001 | 0.31 | -0.32 (0.09) | .0005 | 0.18 | -0.11 (0.03) | .001 | 0.06 |
|  | Concern | -0.98 (0.13) | <.0001 | 0.70 | -1.54 (0.13) | <.0001 | 0.90 | -0.56 (0.05) | <.0001 | 0.33 |
|  | Severity score | -0.86 (0.12) | <.0001 | 0.56 | -0.70 (0.11) | <.0001 | 0.40 | -0.19 (0.04) | <.0001 | 0.09 |

**Notes.** MD = mean difference, SE = standard error, EDE-Q = Eating Disorder Examination Questionnaire, NIAS = Nine Item ARFID Screen, PARDI-AR-Q = PARDI-ARFID-Questionnaire. \* Pairwise comparisons not evaluated for NIAS picky eating, as no significant difference was found in omnibus tests.

**Table 5.**
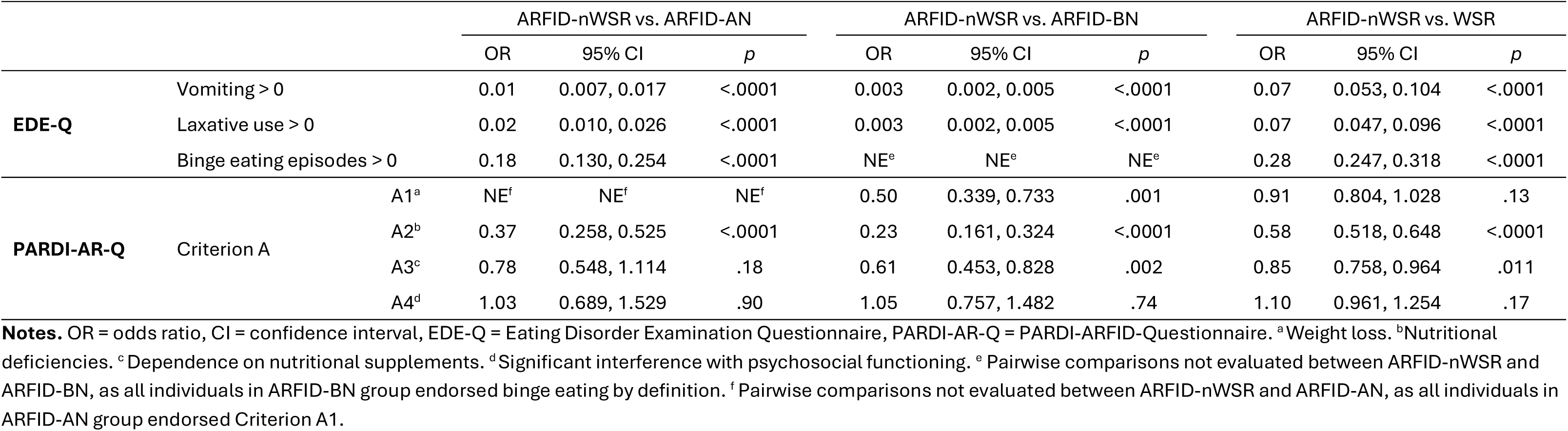
Pairwise comparisons between the ARFID-nWSR and the ARFID-AN, ARFID-BN, and ARFID-WSR (referent groups) for the presence/absence of ED behaviors and PARDI-AR-Q DSM-5 Criteria A1-A4.

### Age and BMI

The mean age of ARFID-nWSR was significantly lower than the mean age of ARFID-BN and ARFID-WSR. We did not find a significant age difference between ARFID-nWSR and ARFID-AN. Regarding BMI, we found ARFID-nWSR to have a significantly higher BMI than ARFID-AN and a significantly lower BMI than ARFID-BN and ARFID-WSR.

### EDE-Q

ARFID-nWSR demonstrated significantly lower EDE-Q scores (global score and subscale scores) compared to all other groups (all *p*s > .0001; Table 4). Additionally, ARFID-nWSR showed significantly lower odds of binge eating than ARFID-AN and ARFID-WSR. Pairwise comparisons for binge eating were not evaluated between ARFID-nWSR and ARFID-BN because all individuals in ARFID-BN endorsed binge eating by definition. Finally, odds of self-induced vomiting and laxative use were lower in ARFID-nWSR than all other groups (Table 5). Our sensitivity analysis excluding the five EDE-Q items that were used for group definition did not change the pattern of our results. ARFID-nWSR demonstrated lower EDE-Q scores and lower odds of binge eating, self-induced vomiting, and laxative use (all *p*s > .0001; results not shown).

### NIAS & PARDI-AR-Q

#### ARFID-nWSR vs. ARFID-AN

ARFID-nWSR had significantly lower scores on the *NIAS Total score*, *NIAS Low appetite*, and *NIAS Fear* than ARFID-AN. We also found lower scores for ARFID-nWSR on *PARDI-AR-Q Lack of interest*, *PARDI-AR-Q Concern*, and *PARDI-AR-Q Severity* than for ARFID-AN. We found no significant difference for *PARDI-AR-Q Sensory*. ARFID-nWSR also had significantly lower odds of meeting PARDI-AR-Q Criterion A2 (nutritional deficiencies) than ARFID-AN. No significant differences were observed between ARFID-nWSR and ARFID-AN for PARDI-AR-Q Criteria A3 (dependence on nutritional supplements) and A4 (significant interference with psychosocial functioning). Pairwise comparisons for PARDI-AR-Q Criterion A1 (weight loss) were not evaluated between ARFID-nWSR and ARFID-AN because all individuals in ARFID-AN endorsed this criterion.

#### ARFID-nWSR vs. ARFID-BN

ARFID-nWSR had significantly lower scores on the *NIAS Total score* and *NIAS Fear* than ARFID-BN. We observed no significant difference for *NIAS Low appetite*. We found higher scores for ARFID-nWSR on *PARDI-AR-Q Sensory,* and lower scores on *PARDI-AR-Q Lack of interest*, *PARDI-AR-Q Concern,* and *PARDI-AR-Q Severity* than for ARFID-BN. Additionally, ARFID-nWSR had significantly lower odds of meeting PARDI-AR-Q Criteria A1, A2, and A3 than ARFID-BN. There was no significant difference between the two groups for PARDI-AR-Q Criterion A4.

#### ARFID-nWSR vs. WSR

ARFID-nWSR had significantly lower scores on the *NIAS Total score* and *NIAS Fear* than ARFID-WSR. We observed no significant difference for *NIAS Low appetite*. We found higher scores for ARFID-nWSR on *PARDI-AR-Q Sensory,* and lower scores on *PARDI-AR-Q Lack of interest*, *PARDI-AR-Q Concern,* and *PARDI-AR-Q Severity* than for ARFID-WSR. ARFID-nWSR had significantly lower odds of meeting PARDI-AR-Q Criteria A2 and A3. We observed no significant difference for PARDI-AR-Q Criteria A1 and A4.

## Discussion

This study is the to date largest investigation of the overlap of ARFID and other ED symptoms in adults with probable ARFID, that is, those who met DSM-5-TR Criterion A on the PARDI-AR-Q and scored above the cutoff on at least one ARFID dimension on the NIAS. We identified those meeting DSM-5-TR Criterion C (ARFID-nWSR) and those who did not (ARFID-AN, ARFID-BN, ARFID-WSR—henceforth referred to as ARFID-ED) by reviewing each question of the EDE-Q under the consideration of potential ARFID-specific interpretations and BMI. We compared ARFID and other ED symptoms across groups and found, as expected, that ARFID-ED reported higher scores on all outcomes assessing non-ARFID ED symptoms (EDE-Q subscales and behaviors). Unexpectedly however, all ARFID dimension scores—except for *NIAS Picky eating* and *PARDI-AR-Q Sensory*—were significantly higher in ARFID-ED than in ARFID-nWSR. Similarly, most physical and psychosocial consequences (PARDI-AR-Q Criteria A1-A4) associated with ARFID were more likely to be endorsed by ARFID-ED than by ARFID-nWSR.

The observed overlap of ARFID and other ED symptoms affords two main interpretations. The first is that the symptoms used to define ARFID are not specific to the condition but rather are transdiagnostic and can be seen in other EDs. Picky eating and low appetite/interest in eating may be endorsed by individuals with other EDs, particularly AN, as these symptoms can result from chronic restriction (5)—a feature shared by ARFID and other EDs. Similarly, fear of aversive consequences of eating is not unique to ARFID; as such consequences, including GI discomfort, are also observed in both AN and BN (31). Furthermore, the physical and psychosocial consequences listed under Criterion A of the DSM-5-TR Criteria lack specificity, as these outcomes are not exclusive to ARFID and are often seen in other EDs as well. If these symptoms are not ARFID-specific, it follows that the instruments used to measure these symptoms—such as the NIAS and PARDI-AR-Q—may not be suitable for differentiating ARFID from other EDs. Previous studies have shown that the NIAS is not suitable for distinguishing ARFID from other EDs when used alone (23, 25), and although Mulkens (16) suggested that the PARDI-AR-Q may be better suited for this purpose, our results do not support this assumption.

The second interpretation is that DSM-5-TR Criterion C may not fully reflect real-world symptomatology, and that ARFID symptoms can, in fact, co-occur with those of other EDs. The possible existence of a mixed phenotype characterized by both ARFID and other ED features is suggested by the fact that the ARFID dimension scores of ARFID-ED in the current study were higher than (and not just equal to) the ARFID dimension scores in clinical and non-clinical samples of other EDs only (23, 25, 26). When comparing the mixed phenotype to an ARFID-only phenotype, participants in this study demonstrated higher scores on most ARFID dimensions and a greater likelihood of most physical and psychosocial consequences in ARFID-ED compared to ARFID-nWSR. This suggests that this mixed presentation is associated with more severe symptomatology. Individuals with this phenotype may experience a “double burden”— experiencing both ARFID-related drivers of restriction and weight- and shape-related concerns. Research and firsthand accounts of co-occurring ARFID and other EDs highlight how the two conditions can exacerbate one another, with symptoms of one potentially worsening or even triggering symptoms of the other (5–10). This aligns with previous findings, indicating a heightened vulnerability to general psychopathology among those with both ARFID and other EDs during their lifetime (32).

The definition of ARFID—particularly clarifying its overlap with and distinctions from other EDs—remains a complex issue that is actively debated. Zickgraf et al. (28) proposed revised diagnostic criteria that formally include the drivers of food avoidance and allow the co-occurrence of ARFID with other EDs. Since then, several researchers have responded to this proposal, either endorsing (33) or rejecting it (34, 35). In particular, Eddy and Negi (33) support revising the DSM-5-TR Criteria for ARFID. However, rather than endorsing a dual diagnosis, they propose a hierarchical dimensional model that captures overlapping and interconnected symptoms, thereby accounting for individual variation in symptom presentation and providing guidance for both clinical care and research. Similarly, while highlighting the value of empirical classification methods in clarifying how ARFID differs from other EDs and other mental health conditions, Richson et al. (36) proposed RDoC (37) and HiTOP (38) as two potential additional frameworks for rethinking ARFID’s classification and therefore shifting the focus from categorical diagnoses to dimensional scores, offering a more nuanced understanding of ARFID within a larger psychopathology landscape.

As requested by Attia and Walsh (34), we provide additional empirical evidence relevant to evaluating the proposed criteria revisions by Zickgraf et al. (28). However, the observed overlap of ARFID and other ED symptoms in these data does not, on its own, support either endorsement or rejection of the proposal. If ARFID dimensions are not unique to ARFID but also present in other EDs (interpretation 1 above), it raises the question whether the inclusion of ARFID dimensions in proposed Criterion A improves diagnostic specificity. In this case, ARFID would continue to be defined by symptoms shared across EDs and remain a diagnosis of exclusion. If the current findings are interpreted as evidence of a mixed phenotype (interpretation 2 above), including the ARFID dimensions in proposed Criterion A could improve diagnostic specificity and justify an ARFID diagnosis alongside other ED symptoms.

### Strengths, Limitations, and Future Directions

A strength of the current study is the large sample drawn from the community. Furthermore, all participants had to meet established criteria on <u>both</u> the NIAS <u>and</u> PARDI-AR-Q, in line with both DSM-5-TR as well as the proposal by Zickgraf et al. (28), thereby enhancing the relevance of our findings to ARFID populations. Additionally, only 5.8% of our sample met screening criteria for AN or BN, indicating our recruitment strategy predominantly targeted individuals with ARFID, rather than a transdiagnostic ED sample. Had this not been the case, we would expect a larger proportion of the sample to meet criteria for AN or BN. Instead, ARFID-nWSR comprised the largest portion of our sample (57.6%). However, we did not identify BED or OSFED in our sample and therefore cannot rule out the possibility that these EDs were present in some individuals in ARFID-WSR and ARFID-nWSR. By focusing on adults, we increased the likelihood of capturing the co-occurrence of ARFID and other ED symptoms, as other EDs typically emerge during adolescence (39). However, our findings are therefore not generalizable to child populations with ARFID. Finally, we used a novel approach to evaluate WSR in ARFID by reviewing each item of the EDE-Q in the context of ARFID and BMI. The significantly lower EDE-Q subscale scores in the ARFID-nWSR group compared to ARFID-ED and the lower odds of all investigated compensatory behaviors in our main, as well as our sensitivity analyses, provide evidence for the effectiveness of our EDE-Q-based criteria for differentiating ARFID *without* other ED psychopathology from ARFID *with* other ED psychopathology.

However, our study also has notable limitations. Our assessment of ARFID relied on self-report measures that addressed DSM-5-TR Criterion A, ARFID dimensions, and Criterion C, but did not capture Criteria B or D (e.g., food availability, cultural practices, or concurrent other conditions). Due to the large sample size, we could not confirm ARFID diagnosis with clinical interviews and thus used the term “probable ARFID” for the entire sample. Finally, while our operationalization of WSR using selected EDE-Q items appears to have been effective by virtue of the significantly lower EDE-Q scores and behaviors in ARFID-nWSR than ARFID-ED in our main and sensitivity analysis, this approach has not been validated.

Extension of this work to clinically diagnosed samples could examine how individuals with ARFID score on non-ARFID ED measures and vice versa. This approach would provide additional insight into the ways ARFID can co-occur with other EDs. However, under current DSM-5-TR criteria, individuals with an ARFID diagnosis are unlikely to endorse other ED symptoms, as Criterion C excludes their co-occurrence. Therefore, large population-based studies that intentionally recruit individuals with any disordered eating behavior are needed. Such samples would likely capture a diverse range of symptom presentations, including individuals who endorse only ARFID symptoms, only non-ARFID ED symptoms, or both. Additionally, large-scale genetic datasets including individuals with ARFID and those with other EDs could help determine whether these conditions—especially ARFID and AN—are genetically separable, offering insights that may not be possible on the phenotypic level alone (40).

### Conclusions

Current limitations of the ARFID diagnosis in the context of other ED behaviors and cognitions have proven challenging for clinicians and researchers, since they may struggle to determine when an ARFID diagnosis is appropriate and encounter difficulties in tailoring treatment to individual symptom patterns (5–7, 41). Although many clinicians are able to navigate the nuanced realities of how ARFID presents in a clinical population—including recognizing the importance of an ARFID diagnosis even with co-occurring weight or shape disturbances—firsthand patient experience suggests this is not always the case (10). Our findings indicate that among those with probable ARFID, those with co-occurring probable AN, probable BN, and WSR may suffer more medical consequences and experience ARFID dimensions more intensely than those with ARFID only. This suggests that, unless treatment of other EDs is adapted to include screening and consideration of ARFID-related restriction motivators, excluding those with co-occurring other ED psychopathology from receiving an ARFID diagnosis may impede those whose ARFID symptoms are most severe from accessing comprehensive treatment that addresses the full range of their ED pathology.

## Summary

- According to DSM-5-TR Criterion C, ARFID cannot be diagnosed alongside anorexia nervosa, bulimia nervosa, or any other body image disturbance, but emerging evidence suggests mixed presentations exist.
- In this study, adults with probable ARFID and other eating disorder pathology reported more severe ARFID symptoms as measured by NIAS and PARDI-AR-Q than those with probable ARFID only.
- DSM-5-TR may not fully capture real-word symptomology, potentially preventing those with mixed presentations of both ARFID and other eating disorder symptoms from receiving care tailored to their complex clinical profiles.

## Public significance statement

Contrary to DSM-5-TR Criteria, this study indicates that ARFID may co-occur with weight- and/or shape-motivated restriction. We found this mixed symptom pattern to be associated with more severe ARFID symptoms and a greater likelihood of physical and psychosocial consequences related to the eating problems. Revising diagnostic criteria may potentially improve outcomes for those with the most severe symptoms by ensuring accurate diagnoses that reflect their clinical presentation and facilitating appropriate care.

## Data Availability

These data that support the findings of this study will be openly available from the National Data Archive (NDA), study C4467.

