## Supplemental material for "Co-occurring weight- and/or shape-motivated restriction in 5,747 adults with probable avoidant/restrictive food intake disorder"

**Table S1.** Review of each item of the Eating Disorder Examination Questionnaire (EDE-Q; Fairburn & Beglin, 2008) items and consideration of possible interpretations in the context of ARFID and BMI.

| Item | Question | Subscale / Behavior | Could be endorsed by someone with ARFID without WSR? | Comment |
|---|---|---|---|---|
| **1** | **Have you been deliberately <u>trying</u> to limit the amount of food you eat to influence your shape or weight (whether or not you have succeeded)?** | **Restraint** | **Depends on BMI** | **For individuals currently or previously at a high BMI, endorsing this item might not necessarily indicate weight- or shape-motivated restriction indicative of an ED cognition other than ARFID. For example, they may be following medical advice to manage their weight.** |
| **2** | **Have you gone for long periods of time (8 waking hours or more) without eating anything at all in order to influence your shape or weight?** | **Restraint** | **No** | **Intentionally going long periods of time without eating anything to influence body shape or weight goes beyond merely limiting calories or selecting healthier foods. Endorsement by individuals at any BMI indicates potentially significant weight- and shape-motivated restriction.** |
| 3 | Have you <u>tried</u> to exclude from your diet any foods that you like in order to influence your shape or weight (whether or not you have succeeded)? | Restraint | Yes | May be endorsed by individuals with ARFID without weight- or shape-motivated restriction indicative of another ED cognition. Those with a currently or previously high BMI may be following medical advice to exclude high-calorie foods, and those with a low BMI may try to exclude low-calorie safe foods to gain weight. |
| 4 | Have you <u>tried</u> to follow definite rules regarding your eating (for example, a calorie limit) in order to influence your shape or weight (whether or not you have succeeded)? | Restraint | Yes | May be endorsed by individuals with ARFID without weight- or shape-motivated restriction indicative of another ED cognition. Those with a currently or previously high BMI may be following medical advice to exclude high-calorie foods, and those with a low BMI may try to exclude low-calorie safe foods to gain weight. |
| **5** | **Have you had a definite desire to have an <u>empty</u> stomach with the aim of influencing your shape or weight?** | **Restraint** | **No** | **Indicates problematic eating disorder behaviors/cognitions regardless of BMI. Intentionally maintaining an empty stomach to influence body shape or weight goes beyond merely limiting calories or selecting healthier foods.** |
| 6 | Have you had a definite desire to have a <u>totally flat</u> stomach? | Shape concern | Yes | May be endorsed by individuals with ARFID who have specific fears of gastrointestinal symptoms (e.g., painful bloating) and not necessarily indicate a weight or shape disturbance. |
| 7 | Has thinking about <u>food, eating, or calories</u> made it very difficult to concentrate on things you are interested in (for example, working, following a conversation, or reading)? | Eating concern | Yes | Could reasonably indicate an ARFID cognition (e.g., ruminating about availability of safe foods). |

| | | | | |
|---|---|---|---|---|
| 8 | Has thinking about shape or weight made it very difficult to concentrate on things you are interested in (for example, working, following a conversation, or reading)? | Weight concern/Shape concern | Yes | May be endorsed by individuals with ARFID without weight- or shape-motivated restriction indicative of another ED cognition. Those with a currently or previously high BMI may worry about being overweight, while those with a low BMI may worry about being underweight. Individuals with normal BMIs may endorse this item due to a past history of being over or underweight and/or based on cultural messaging about weight and shape. |
| 9 | Have you had a definite fear of losing control over eating? | Eating concern | Yes | May be endorsed by individuals with ARFID without weight- or shape-motivated restriction indicative of another ED cognition. For example, meeting nutritional requirements may require a high level of control, and losing control could mean not eating enough. Additionally, binge eating is not an exclusion criterion for ARFID. Individuals with ARFID may experience binges with loss of control after not eating for long periods of time due to psychosocial concern and/or lack of access to safe foods. |
| 10 | Have you had a definite fear that you might gain weight? | Shape concern | Yes | May be endorsed by individuals with ARFID and a currently or previously high BMI, e.g., following medical advice to manage their weight or due to cultural messaging about weight and shape. |
| **11** | **Have you felt fat?** | **Shape concern** | **Depends on BMI** | **Item may be endorsed by individuals with ARFID with a currently or previously high BMI and those with "normal" BMIs based on cultural messaging about weight and shape.** |
| **12** | **Have you had a strong desire to lose weight?** | **Weight concern** | **Depends on BMI** | **Item may be endorsed by individuals with ARFID and a currently or previously high BMI e.g., following medical advice to manage their weight or due to cultural messaging about weight and shape.** |
| 13 | Over the past 28 days, how many times have you eaten what other people would regards as an unusually large amount of food (given the circumstances)? | Binge eating | Yes | Binge eating is not an exclusion criterion for ARFID. Individuals with ARFID may experience binges with loss of control after not eating for long periods of time due to psychosocial concern and/or lack of access to safe foods. |
| 14 | ...On how many of these times did you have a sense of having lost control over your eating (at the time you were eating)? | Binge eating | Yes | Binge eating is not an exclusion criterion for ARFID. Individuals with ARFID may experience binges with loss of control after not eating for long periods of time due to psychosocial concern and/or lack of access to safe foods. |

| | | | | |
|---|---|---|---|---|
| 15 | Over the past 28 days, how many days have such episodes of overeating occurred (i.e., you have eaten an unusually large amount of food and have had a sense of loss of control at the time)? | Binge eating | Yes | Binge eating is not an exclusion criterion for ARFID. Individuals with ARFID may experience binges with loss of control after not eating for long periods of time due to psychosocial concern and/or lack of access to safe foods. |
| 16 | Over the past 28 days, how many times have you made yourself sick (vomit) as a means of controlling your shape or weight? | Self-induced vomiting | Yes | According to DSM-5, self-induced vomiting is not an exclusion criterion for ARFID. |
| 17 | Over the past 28 days, how many times have you taken laxatives as a means of controlling your shape or weight? | Laxative use | Yes | According to DSM-5, laxative use is not an exclusion criterion for ARFID. |
| 18 | Over the past 28 days, how many times have you exercised in a "driven" or "compulsive" way as a means of controlling your weight, shape, or amount of fat, or to burn off calories? | Excessive exercise | Yes | Item may be endorsed by individuals with ARFID with a currently or previously high BMI who, for example, may be following medical advice to manage their weight through exercise. |
| 19 | Over the past 28 days, on how many days have you eaten in secret (i.e., furtively)? Do not count episodes of binge eating. | Eating concern | Yes | Item may be endorsed by individuals with ARFID and specific food preferences. Individuals with ARFID may feel embarrassed about eating the same foods repeatedly and worry about being judged. |
| 20 | On what proportion of the times that you have eaten have you felt guilty (felt that you have done wrong) because of its effect on your shape or weight? ... Do not count episodes of binge eating. | Eating concern | Yes | Item may be endorsed by individuals with ARFID and specific food preferences. Individuals with ARFID may feel guilty about eating the same foods repeatedly. |
| 21 | Over the past 28 days, how concerned have you been about other people seeing you eat? ... Do not count episodes of binge eating. | Eating concern | Yes | Item may be endorsed by individuals with ARFID and specific food preferences. Individuals with ARFID may feel embarrassed about eating the same foods repeatedly and worry about being judged. |
| 22 | Over the past 28 days, has your weight influenced how you think about (judge) yourself as a person? | Weight concern | Yes | Those with a currently or previously high BMI may worry about being overweight and those with a low BMI may worry about being underweight. Those with "normal" BMIs may endorse this item based on cultural messaging about weight. |
| 23 | Over the past 28 days, has your shape influenced how you think about (judge) yourself as a person? | Shape concern | Yes | Those with a currently or previously high BMI may worry about being overweight and those with a low BMI may worry about being underweight. Those with "normal" BMIs may endorse this item based on cultural messaging about shape. |

| | | | | |
|---|---|---|---|---|
| 24 | Over the past 28 days, how much would it have upset you if you had been asked to weigh yourself once a week (no more, or less, often) for the next four weeks? | Weight concern | Yes | Those with a currently or previously high BMI may worry about being overweight and those with a low BMI may worry about being underweight. Those with "normal" BMIs may endorse this item based on cultural messaging about weight. |
| 25 | Over the past 28 days, how dissatisfied have you been with your weight? | Weight concern | Yes | Those with a currently or previously high BMI may worry about being overweight and those with a low BMI may worry about being underweight. Those with "normal" BMIs may endorse this item based on cultural messaging about weight. |
| 26 | Over the past 28 days, how dissatisfied have you been with your shape? | Shape concern | Yes | Those with a currently or previously high BMI may worry about being overweight and those with a low BMI may worry about being underweight. Those with "normal" BMIs may endorse this item based on cultural messaging about shape. |
| 27 | Over the past 28 days, how uncomfortable have you felt seeing your body (for example, seeing your shape in the mirror, in a shop window reflection, while undressing or taking a bath or shower)? | Shape concern | Yes | Those with a currently or previously high BMI may worry about being overweight and those with a low BMI may worry about being underweight. Those with "normal" BMIs may endorse this item based on cultural messaging about shape. |
| 28 | Over the past 28 days, how uncomfortable have you felt about others seeing your shape or figure (for example, in communal changing rooms, when swimming, or wearing tight clothes)? | Shape concern | Yes | Those with a currently or previously high BMI may worry about being overweight and those with a low BMI may worry about being underweight. Those with "normal" BMIs may endorse this item based on cultural messaging about shape. |

**Notes.** WSR = Weight- or shape- motivated restriction. Bolded items were chosen for the identification of WSR.

**Table S2.** Pearson correlations of age, BMI, EDE-Q, NIAS, and PARDI-AR-Q variables

| | | | | EDE-Q | | | | | NIAS | | | | PARDI-AR-Q | | | |
|---|---|---|---|---|---|---|---|---|---|---|---|---|---|---|---|---|
| | | Age | BMI | GS | R | EC | WC | SC | TS | PE | LA | F | S | LI | C | SV |
|  | Age | ---- |  |  |  |  |  |  |  |  |  |  |  |  |  |  |
|  | BMI | 0.09 | ---- |  |  |  |  |  |  |  |  |  |  |  |  |  |
| EDE-Q | GS | 0.14 | 0.26 | ---- |  |  |  |  |  |  |  |  |  |  |  |  |
|  | R | 0.20 | 0.14 | 0.89 | ---- |  |  |  |  |  |  |  |  |  |  |  |
|  | EC | 0.04 | 0.18 | 0.87 | 0.72 | ---- |  |  |  |  |  |  |  |  |  |  |
|  | WC | 0.12 | 0.32 | 0.94 | 0.75 | 0.75 | ---- |  |  |  |  |  |  |  |  |  |
|  | SC | 0.15 | 0.31 | 0.94 | 0.75 | 0.74 | 0.93 | ---- |  |  |  |  |  |  |  |  |
| NIAS | TS | -0.03 | -0.16 | 0.04 | 0.05 | 0.13 | 0.00 | -0.01 | ---- |  |  |  |  |  |  |  |
|  | PE | -0.09 | 0.07 | 0.04 | 0.00 | 0.07 | 0.03 | 0.03 | 0.40 | ---- |  |  |  |  |  |  |
|  | LA | -0.09 | -0.3 | -0.12 | -0.07 | -0.03 | -0.14 | -0.13 | 0.76 | 0.03 | ---- |  |  |  |  |  |
|  | F | 0.07 | -0.05 | 0.14 | 0.14 | 0.19 | 0.09 | 0.09 | 0.78 | -0.02 | 0.41 | ---- |  |  |  |  |
| PARDI-AR-Q | S | -0.23 | 0.10 | 0.09 | 0.02 | 0.18 | 0.08 | 0.09 | 0.40 | 0.45 | 0.22 | 0.20 | ---- |  |  |  |
|  | LI | -0.10 | -0.16 | 0.00 | 0.01 | 0.07 | -0.03 | -0.02 | 0.62 | 0.08 | 0.72 | 0.38 | 0.29 | ---- |  |  |
|  | C | 0.09 | -0.02 | 0.21 | 0.21 | 0.31 | 0.17 | 0.15 | 0.58 | 0.05 | 0.29 | 0.71 | 0.28 | 0.33 | ---- |  |
|  | SV | -0.14 | 0.02 | 0.10 | 0.10 | 0.27 | 0.13 | 0.13 | 0.36 | 0.34 | 0.18 | 0.24 | 0.44 | 0.21 | 0.40 | ---- |

**Notes.** EDE-Q = Eating Disorder Examination Questionnaire, NIAS = Nine Item ARFID Screen, PARDI-AR-Q = PARDI-ARFID-Questionnaire, GS = global score, R = restraint, EC = eating concern, SC = shape concern, WC = weight concern, TS = total score, PE = picky eating, LA = low appetite, F = fear, S = sensory, LI = low interest, C = concern, SV = severity. Green cells highlight correlations among EDE-Q, NIAS, and PARDI-AR-Q subscale scores.
